# Comprehensive analysis of primary immune thrombocytopenia clinical trials in the ClinicalTrials.gov database

**DOI:** 10.1101/2023.01.16.23284594

**Authors:** Yacan Wang, Hui Bi, Wen Yang, Honghui Wang, Xiuli Wang, Dongmei Xie, Huiting Li, Zeping Zhou

## Abstract

**Objective:** The purpose is to clarify the overall situation of clinical related to primary immune thrombocytopenia (ITP), to evaluate the difference between published clinical trials and unpublished trials, and to evaluate the relevant information of trial publication.

**Methods:** Search the ClinicalTrials.gov database on March 20, 2022 to identify ITP clinical trials and obtain relevant data. Publications in PubMed were searched using standardized methods to identify the publication of completed clinical trials.

**Results:** Of 341 trials identified, interventional trials were the most common (74.2%, n=253). Interventional trials and observational trials differ in the main research content (odds ratio (OR) 0.06, 95% confidence interval (CI) 0.03-0.015, P=0.000). In terms of published articles, there are few trials involving non drugs (OR 0.23, CI 0.08-0.64, P = 0.005). There were fewer trials with less than 10 participants (OR 1.52, CI 1.06-2.20, P = 0.024). Of the 167 completed trials, 93 (55.7%) were published and 48 (28.7%) uploaded results.

**Conclusion:** If the main research content involves drugs, trials with a larger number of people are more likely to be published. The publication rate of ITP clinical trials is high, but the submission rate of database results is low. Therefore, more attention should be paid to the submission of clinical trial results in the later stage.

## Introduction

Primary immune thrombocytopenia (ITP) manifests as peripheral platelets less than 100×10^9^/L, and other causes of thrombocytopenia should be excluded. This disease, also known as idiopathic thrombocytopenic purpura, is an acquired disease that is immunologically mediated.[1] The pathogenesis has not been fully elucidated. There is currently no ‘gold standard’ for this disease, and the diagnosis method is still exclusive. In the past decade or so, great changes have taken place in its second-line treatment. The efficacy of rituximab, thrombopoietin receptor agonists (eltrombopag, avatrombopag, romiplostim), fostamatinib and other drugs have been discovered and are recommended for persistent and chronic ITP.[2, 3] The use of splenectomy has declined following these drug treatments.[4] However, current second-line treatments still suffer from poor longer term outcomes and high drug prices.[5, 6]

ClinicalTrials.gov is a database of clinical trials conducted around the world, already involving 221 countries. ‘Food and Drug Administration Modernization Act of 1997’, a US federal law, requires the registration of efficacy trials of investigational new drugs for serious diseases. In 2000, ClinicalTrials.gov, a website jointly developed by the National Institutes of Health (NIH) and Food and Drug Administration (FDA), was officially opened. In the past 20 years, the scope of clinical trial registration policy has changed many times. Publication policy initiated by the International Committee of Medical Journal Editors (ICMJE) in 2004, with a scope of all interventional trials, including Phase 1 trials. After that, the European Union, the NIH, and the World Medical Association DECLARATION OF HELSINKI have published or revised the corresponding regulations. ‘The Section 801 of the Food and Drug Administration Amendments Act (FDAAA 801)’, which took effect on September 27, 2007, and ‘the Final Rule for Clinical Trials Registration and Results Information Submission’, which took effect on January 18, 2017, set the same requirements for the registration scope of clinical trials: clinical trials of a FDA regulated drug, biological, or device product other than Phase 1 (drug/biological products) or small feasibility studies (device products). In conclusion, all research intended for publication should be registered.

FDA requires that clinical trials be registered at least 21 days after the first subject is recruited. Both ICMJE and Chinese Clinical Trial Registry require and recommend registration before subject recruitment. Researchers will use the ICMJE criteria to assess the timing of trial registration and trial recruitment. FDAAA 801 requires completed trials to upload their results to the ClinicalTrials.gov database within one year. The ClinicalTrials.gov results database was launched in September 2008.

We will analyze the ITP related data in the ClinicalTrials.gov database to understand (1) the characteristics of all ITP clinical trials, (2) the characteristics of interventional and observational trials, (3) the difference between published and unpublished trials, (4) Publication rate, number of published articles, time of publication of articles, publicity of results, new drugs, etc.

## Methods

### Data collection in ClinicalTrials.gov

On March 20, 2022, the researchers searched the ClinicalTrials.gov database for the following terms ‘ITP’, ‘Purpura, Thrombocytopenic, Idiopathic’, ‘Immune Thrombocytopenia’, ‘Primary Immune Thrombocytopenia’. 346, 343, 330, and 69 trials were retrieved, respectively. The information from these trials was then exported to a spreadsheet through the website’s own functionality. The researchers screened all trials, removed duplicate trials, and removed trials not related to the disease, such as drug immune thrombocytopenia, thrombotic thrombocytopenic purpura, and bladder cancer chemotherapy tip regimens. 341 trials were ultimately identified.

### Data Extraction

Extract relevant data from spreadsheets: NCT number, Status, Trial Results, Locations, Age, Phases, Enrollment, Funder, Study Type, Study Designs, First Posted Date, Start Date, Completion Date, Results First Posted. And extract relevant information from the specific website of each trial or the official website of the company: main research content, reasons for not conducting research, type of intervention, type of interventional drug, interventional drug trial study design (single-arm, multi-arm non-random non-double-blind, randomized non-double-blind, double-blind no placebo, double-blind placebo), relevant countries, etc.

### Clarify the publication status of completed trials

Standardized methods were used to search for published articles in clinical trials. First of all, the researchers have obtained relevant data of completed clinical trials through the ClinicalTrials.gov database. Afterwards, the NCT number will be searched on the PubMed website to clarify the publication status of the trial. For trials with no results in searching NCT numbers, PubMed will be searched by disease name, drug name, region, Principal Investigator, and the institution where the Principal Investigator is located. The research type (single-arm or double-blind), phases, study type (interventional or observational), specific study group and other information of the retrieved articles were checked and matched with the information in the ClinicalTrials.gov database. If multiple articles are published in a trial, the oldest article that best matches the trial content will be used. Other researchers will recheck the clinical trial information with the published article information. Finally, determine whether each completed trial has been published, as well as the number of published articles and the time of publication.

### Data analysis

First, the data types are divided. Divide Funder into ‘Industry’, ‘University or hospital or NIH’ and ‘Cooperative sponsorship’. The main content of the trial was divided into medication related, pathogenic, and others (including diagnosis, database, bleeding risk, quality of life, epidemiology, thrombosis risk, surgery). The number of people included in the trial was divided into five intervals. Comparing the start date and the first posted date, trials were divided into ‘register first’ trials and ‘recruit first’ trials. It is divided into four intervals based on ‘Completion Date’ data from completed trials. Finally, binary logistic regression will be used to evaluate factors related to whether the trial can publish articles, as well as the differences between interventional and observational trials. Researchers will provide P values, odds ratios (OR), and 95% confidence intervals (CI).

Researchers used descriptive analysis to assess trial duration, time to publication, and time to submit results after trial completion. First draw the frequency histogram and calculate the relevant data according to the distribution type. Symmetric distribution calculates mean and standard deviation; skewed distribution calculates median and interquartile range. Trials with missing data will be excluded. Analyses were performed using IBM SPSS software (version 9.4; IBM Corp., Armonk, NY, USA).

## Results

### Clinical trial features

Of the 341 trials, interventional trials were the most common type, accounting for 74.2% of all trials (n=253). There were also 86 observational trials, and 2 ‘Expanded Access’ trials. Overall data after excluding 2 expanded access trials are shown in Table 1.

**Table 1.**
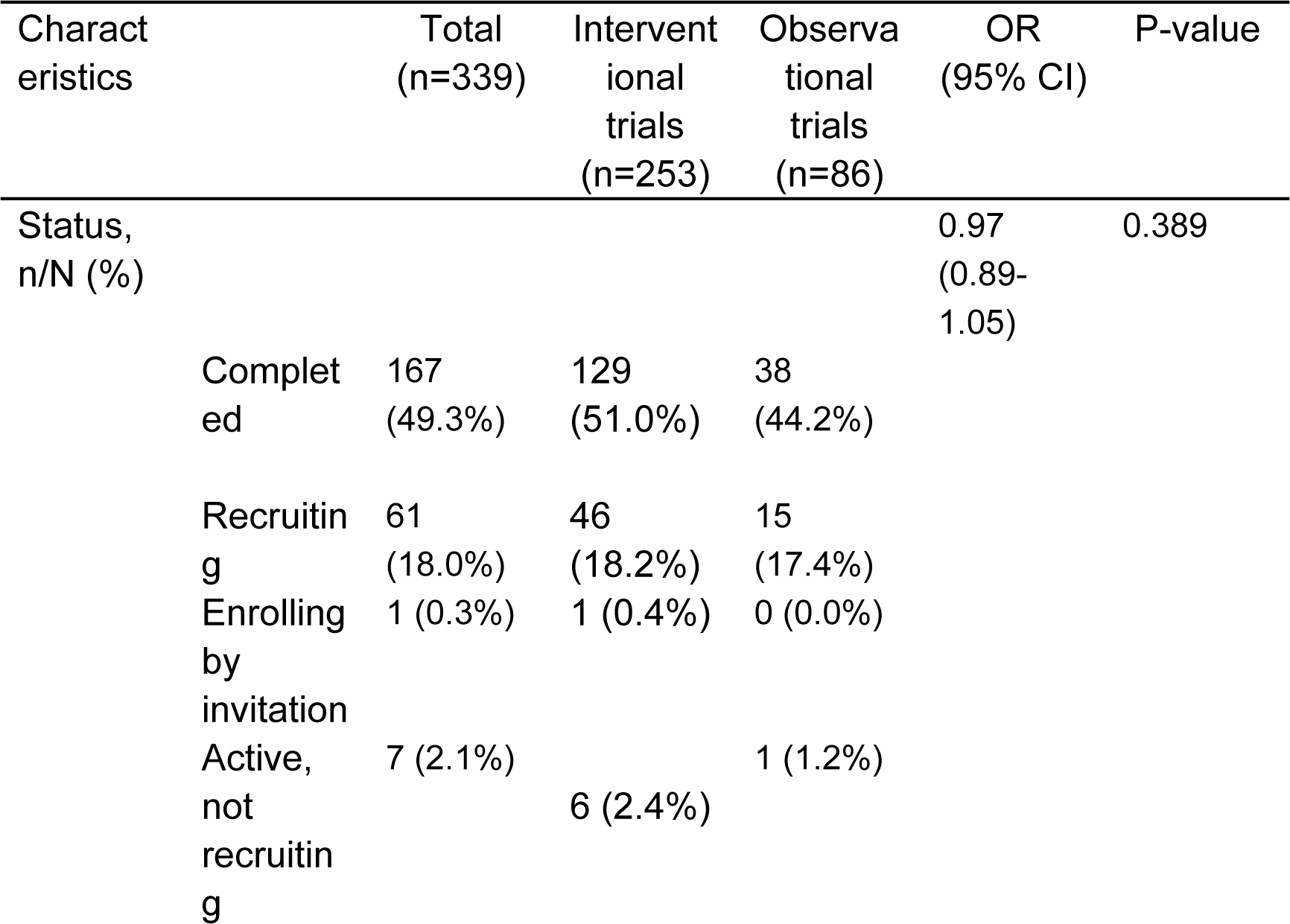
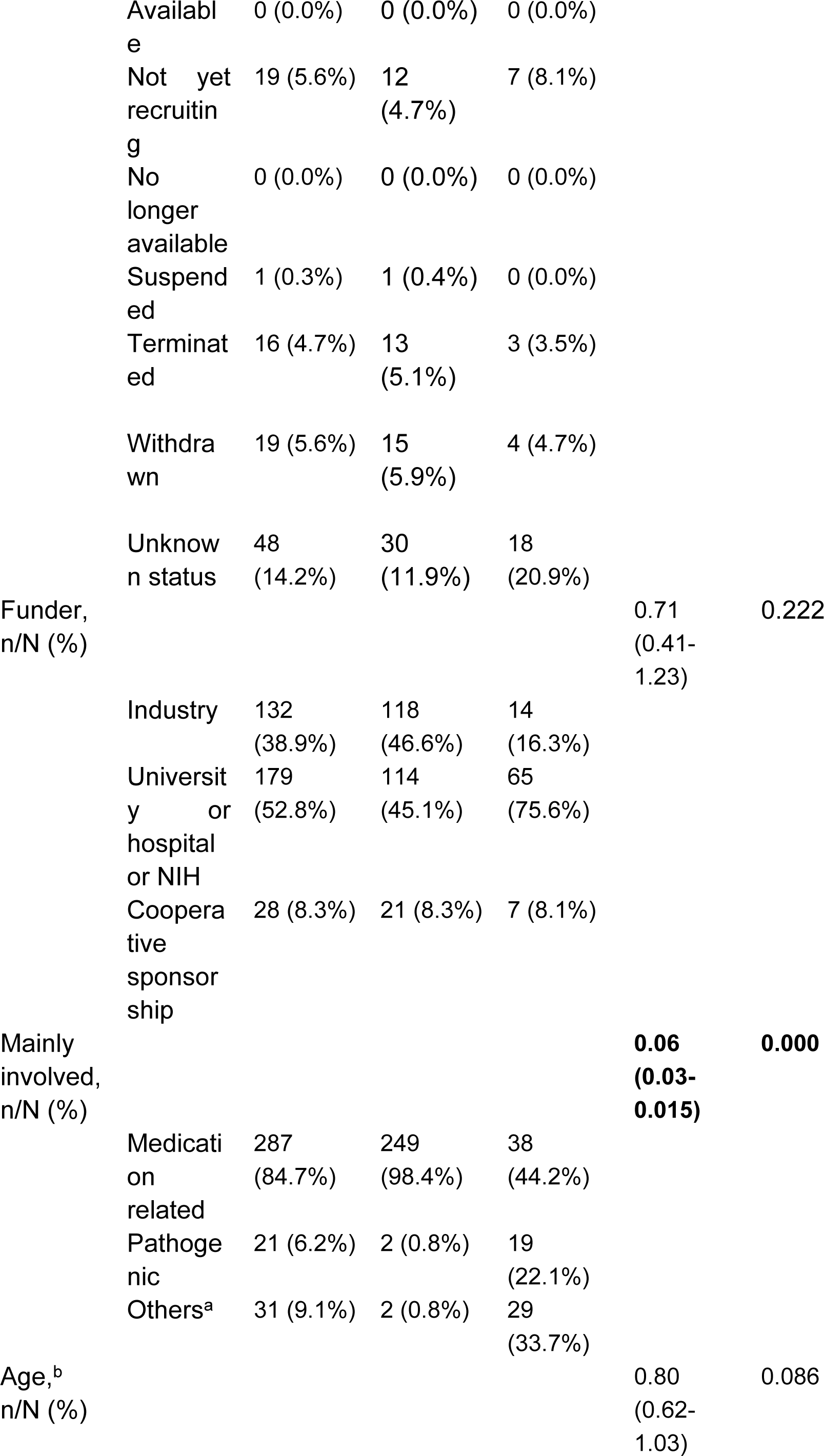
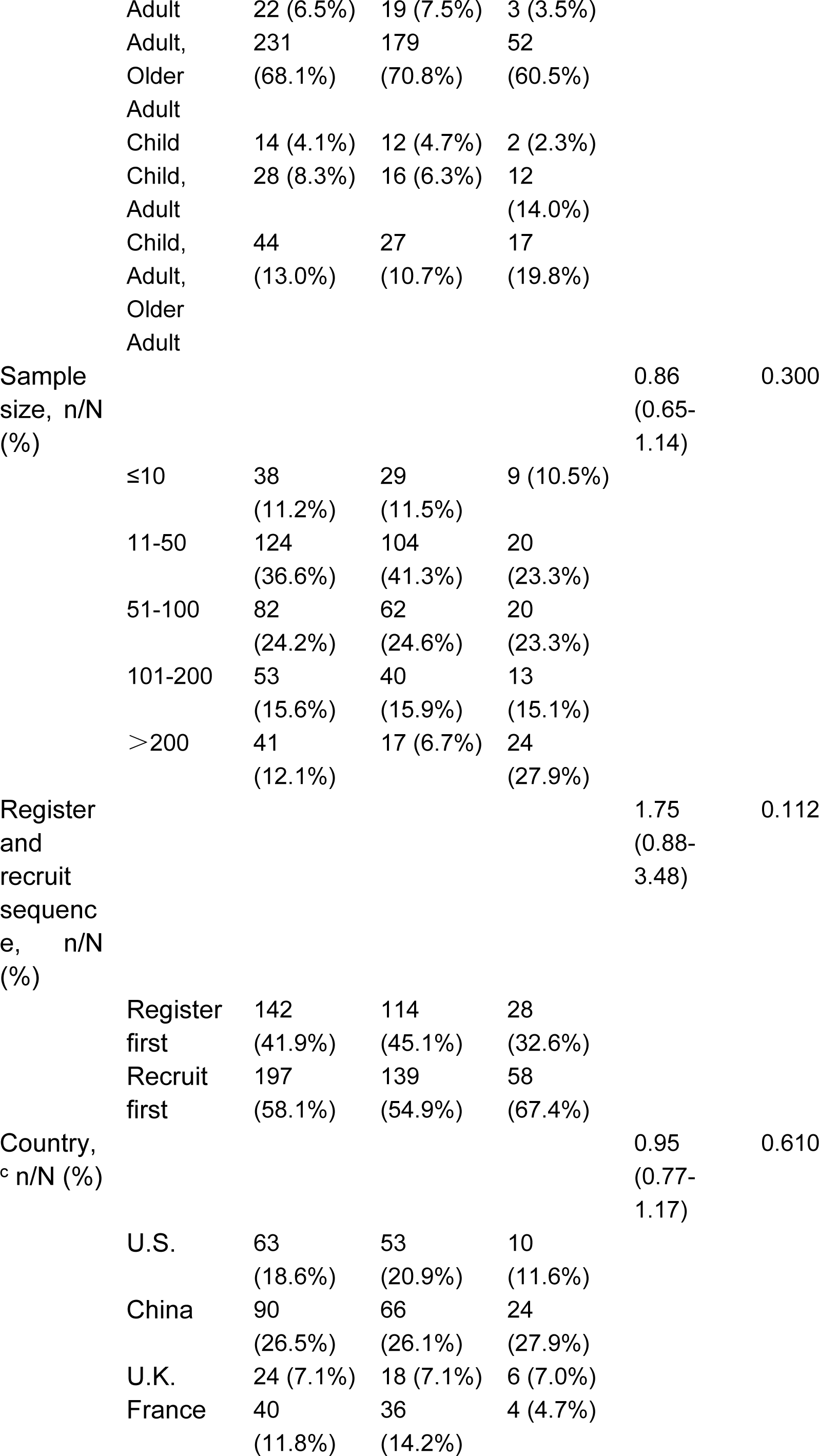
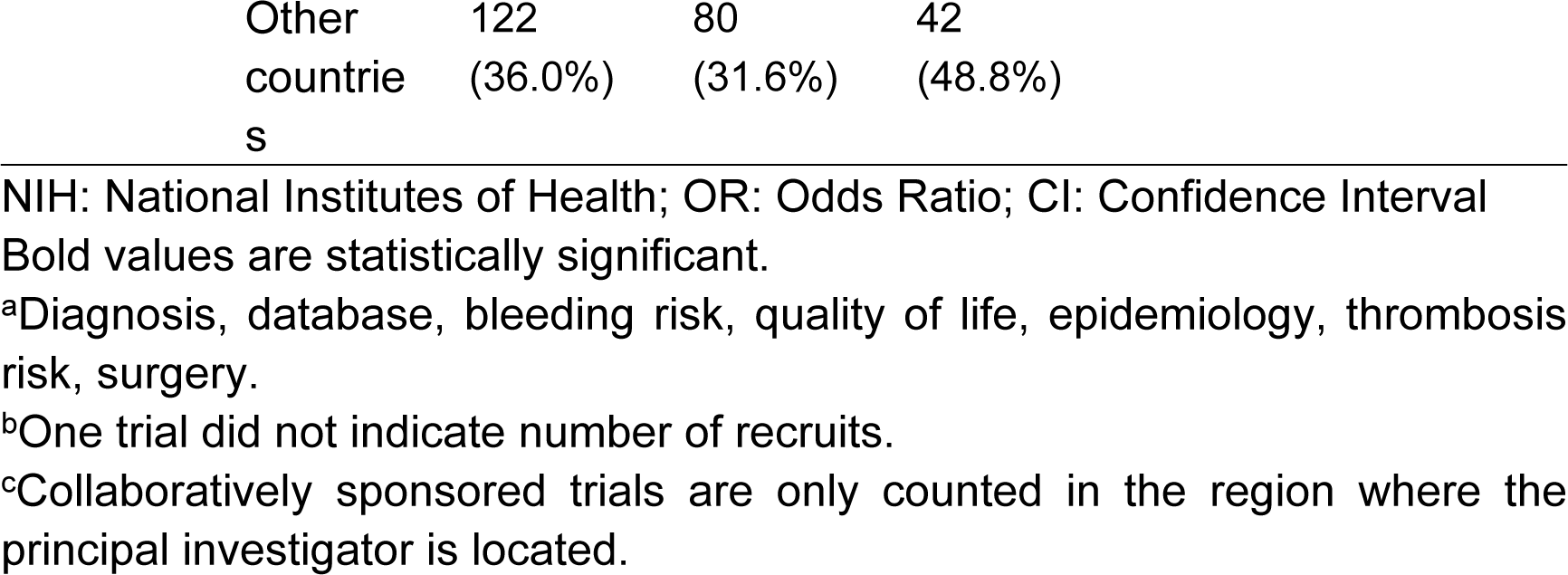
Trends between the two trial types registered on ClinicalTrials.gov

There are 61 trials in ‘Recruiting’ status. In terms of funding, 59.0% are ‘University or Hospital or NIH’ and 34.4% are ‘Industry’. 55.0% of trials funded by ‘University or Hospital or NIH’ were located in China and 17.5% in France. There are 19 trials in the ‘Not yet recruiting’ status. In terms of funding, 78.9% are ‘University or Hospital or NIH’ and 15.8% are ‘Industry’. 56.3% of trials funded by ‘University or Hospital’ were located in China. Seven trials have a status of ‘Active, not recruiting’. 2 trials have uploaded results to the database without updating the status.

When the status of the trial is ongoing, it has passed its estimated completion time, and the data has not been updated for 2 years, the ClinicalTrials.gov database will automatically change the status to ‘Unknown status’.14.2% of the trials showed ‘Unknown status’ (n=48). 83.3% were funded by ‘University or Hospital or NIH’ and 6.3% by Industry. Seven trials published papers, but none were updated with trial status and results.

The ‘Withdrawn’ state indicates that the trial was stopped early before the first participant was enrolled. There are 19 trials, with funding 63.2% ‘University or Hospital or NIH’ and 26.3% ‘Industry’. Of these, 12 trials indicated reasons for withdrawal (seven because of no patient enrollment, and five because of the company’s decision to cancel the trial), and seven for no reason. One of the trials NCT01864512 has published articles.

The ‘Terminated’ status indicates that the trial has been stopped prematurely and will not be restarted. There were 16 trials, 62.5% of which were Funded by industry and 25.0% by ‘University or Hospital or NIH’. Reasons for termination were noted in 11 trials (four trials indicated difficulty in recruiting patients, seven for other reasons such as sponsor decision, need for higher doses of drug, sufficient data collected), and 5 trials did not indicate reasons. Four trials published results on the website. Among them, NCT00225875 has published articles.

### Drug and time relationship

The ICMJE requires trials to be registered before recruiting patients, so the time data are ’the First Posted’ and include the drug class (Fig 1). Since 2005, the number of trials has gradually increased. In the past five years, the number of trials on eltrombopag and rituximab has begun to decline, and the number of new drugs and new therapies has begun to increase.

**Fig 1.**
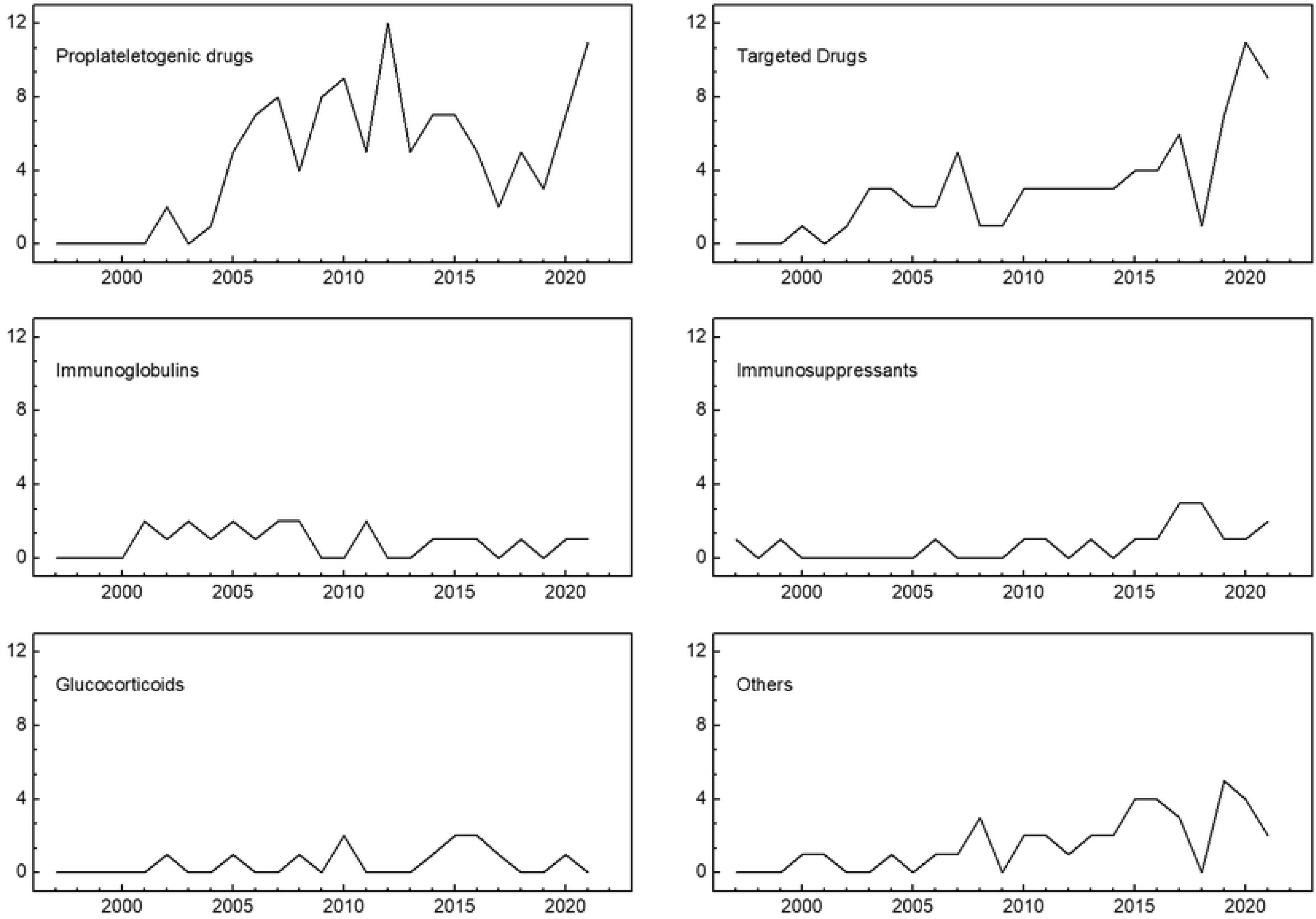
Variation in the number of trials with time for different drugs Some of the trials involve multiple drugs, and the types of main study drugs will be counted.

### Country distribution

Fig 2 shows the number of countries involved in the trials, which have divided funders into ’Industry’ and ’Other’. This found that the total number of trials involving China was the largest, but 85.9% of the trials were conducted by ’University or Hospital’. The total number of trials involving the United States is second, of which the number of ‘Industry’ accounts for 59.7%.

**Fig 2.**
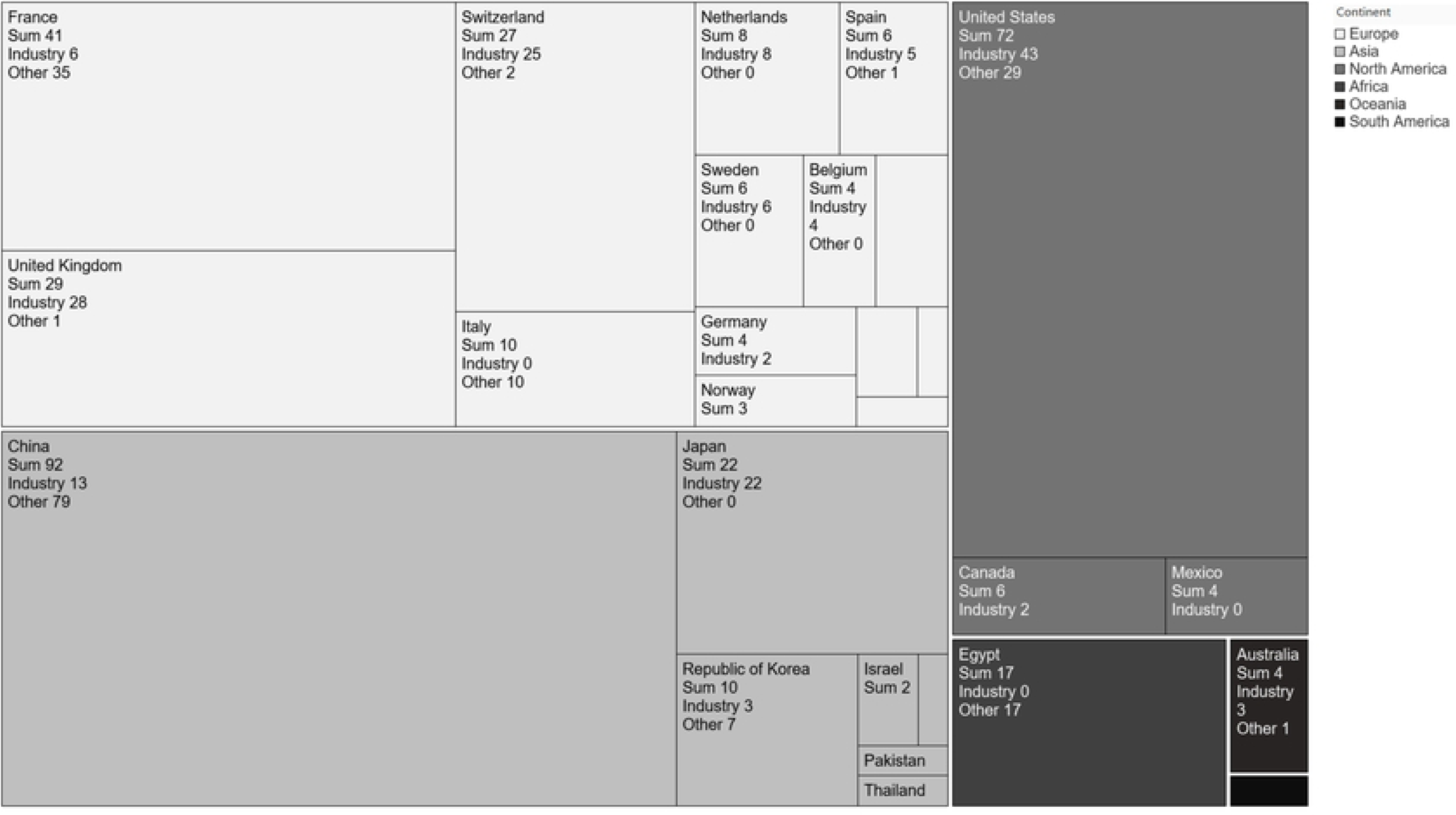
Distribution of countries involved in ITP trials. ‘Other’ means university or hospital funded. Information on the location of cooperatively sponsored trials will be included for all institutions.

### Completed Research Features

There were 167 completed status trials. The median trial duration was 29.2 months (IQR 31.7 months, n=162). The median time to publication after the trial was completed was 24.2 months (IQR 26.2 months, n=93). The median time to submission of results after trial completion was 23.8 months (IQR 35.5 months, n=48). Of the trials funded by Industry, 27.1% were located in the US, 24.7% in the UK, 16.5% in Japan and 11.8% in Switzerland. Of the trials funded by ‘University or Hospital’, 30.8% were located in China, 24.6% in France, 12.3% in the United States, and 9.2% in Italy.

A total of 112 articles were published in 93 (55.7%) trials, 38 (40.9%) had updated results on the website, and one trial had updated results on the website but was not reviewed by the National Library of Medicine. Of the 74 trials unpublished, 10 (13.5%) had their results updated on the website and one was not reviewed. There were 94 completed clinical trials between October 1, 2008, and March 20, 2021, of which 20 (21.3%) had updated results and 6 (6.4%) had results updated within one year.

The differences between the published article trials and the unpublished article trials are shown in Table 2. The main research content was pathogenic or other less likely to be published (OR 0.23, CI 0.08-0.64, P = 0.005). Trials with larger numbers of recruits are easier to publish (OR 1.52, CI 1.06-2.20, P = 0.024). Fewer trials were published in later years (OR 0.56, CI 0.35-0.92, P = 0.021); more trials were published with results uploaded (OR 3.32, CI 1.30-8.48, P = 0.012).

**Table 2.**
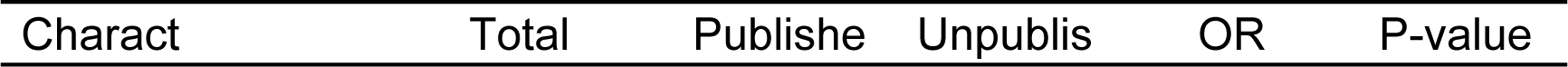
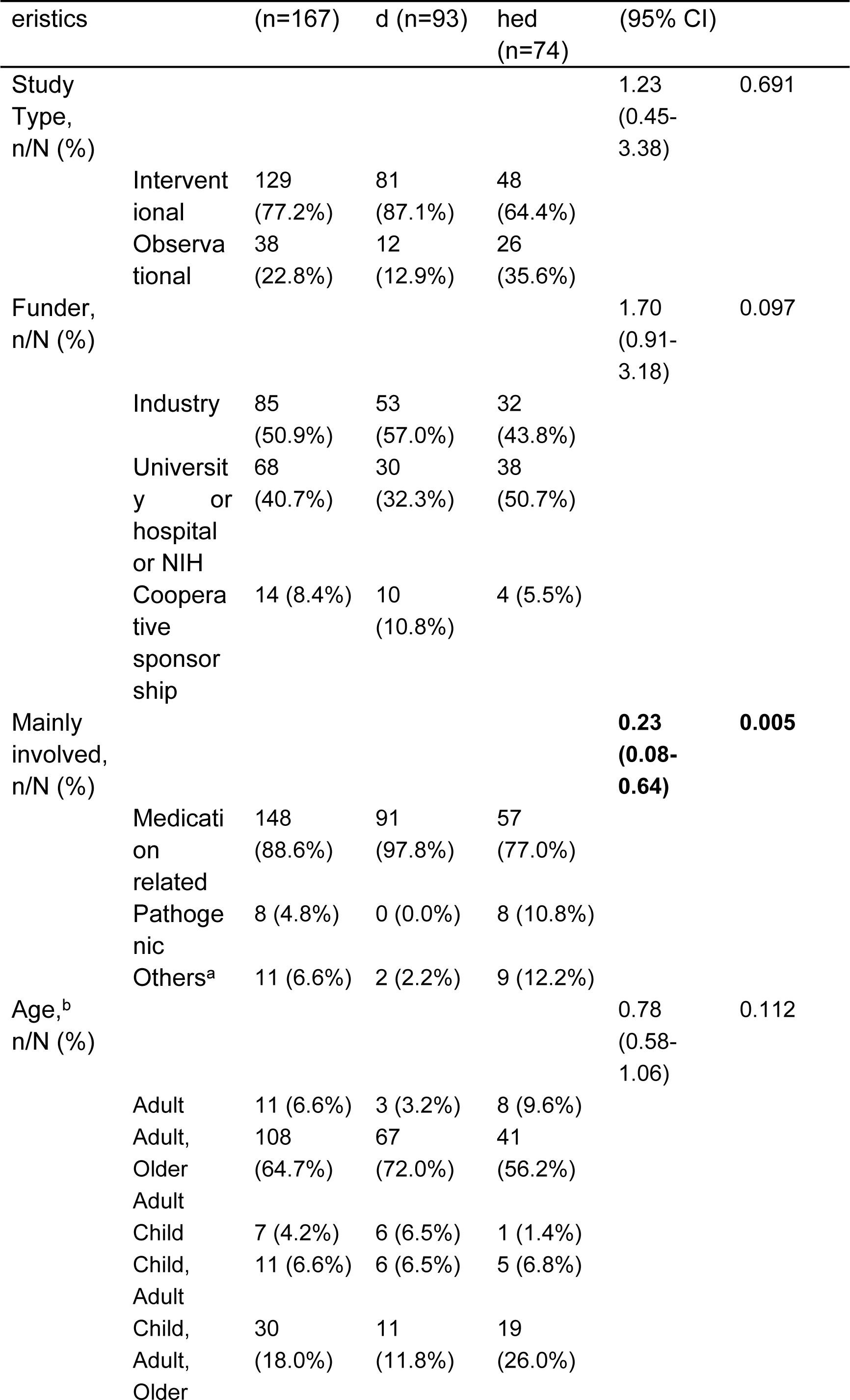
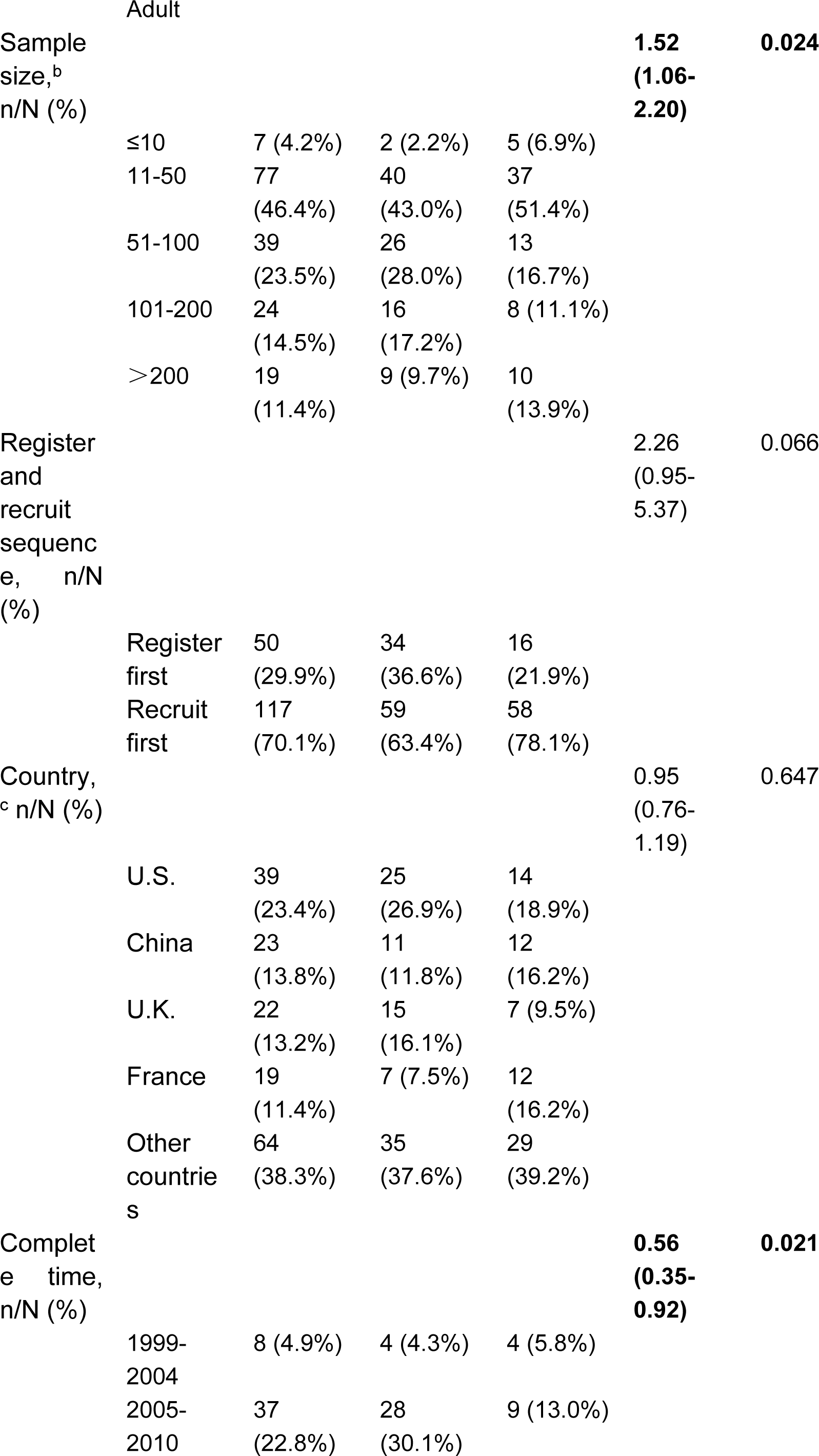
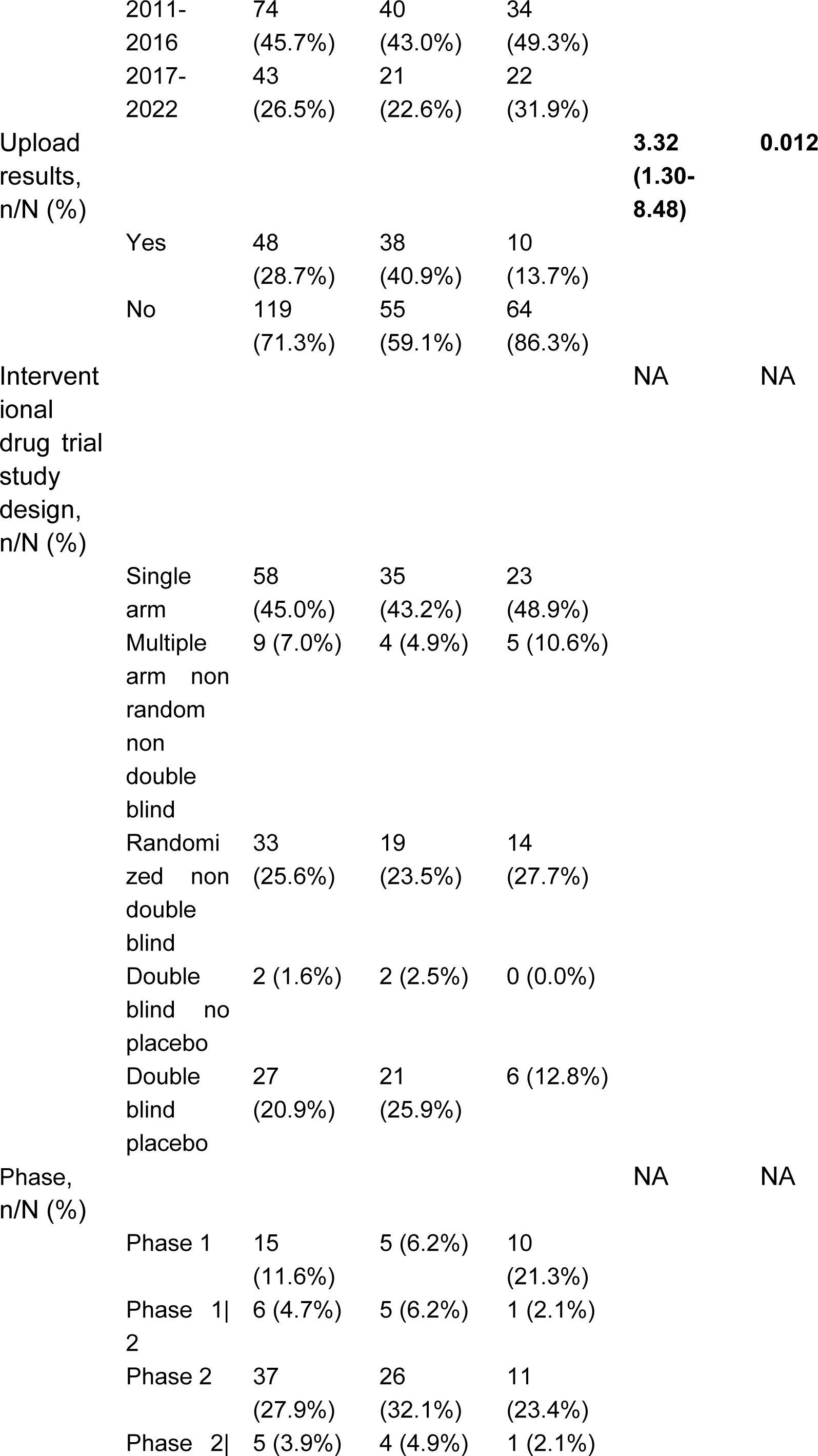
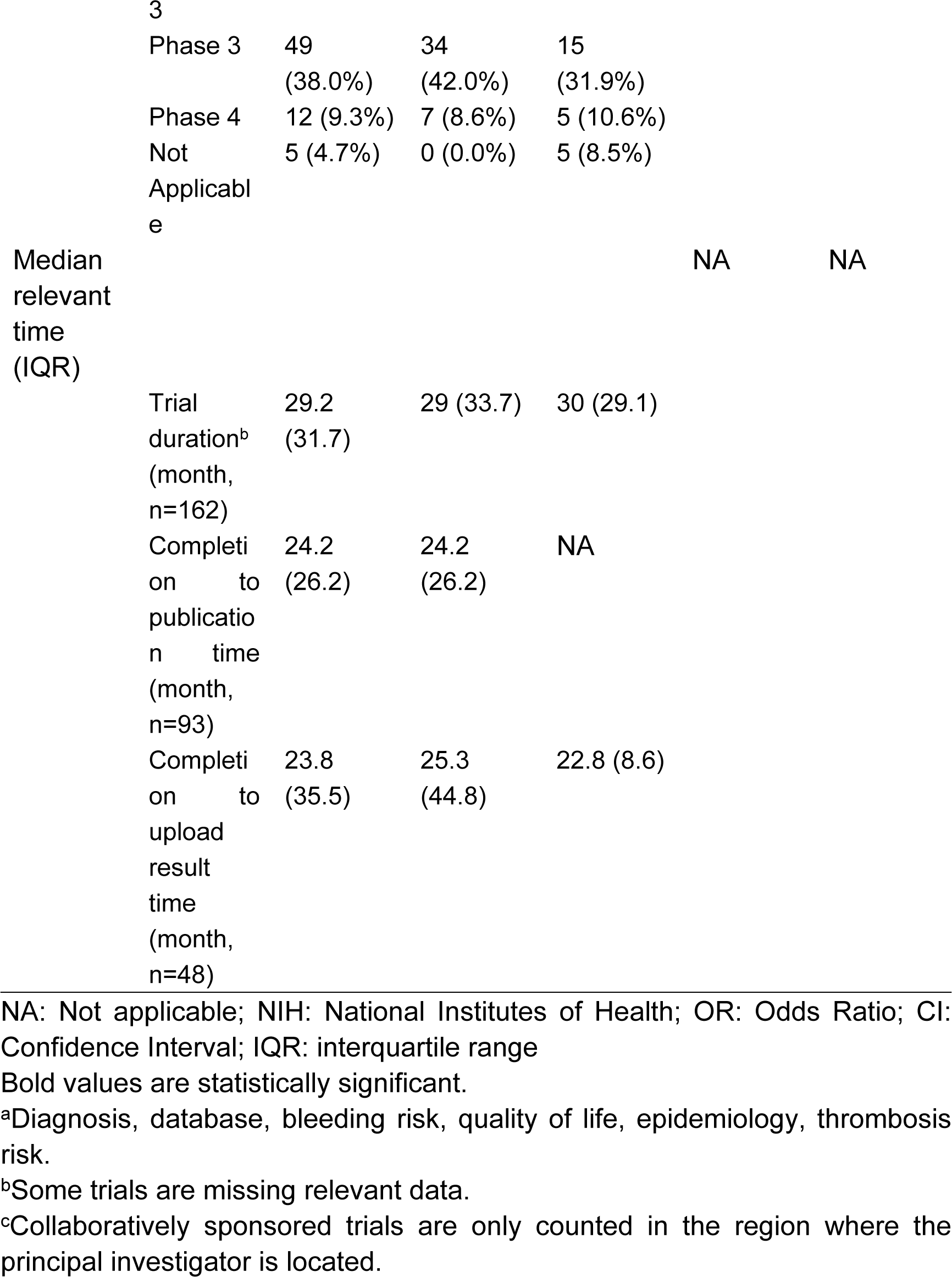
Impact of different characteristics on publication of completed trials registered on ClinicalTrials.gov

### Difference Between Interventional and Observational Trials

See Table 1 for the difference between interventional and observational. In terms of research content, there were more medication related interventional trials than observational trials (OR 0.05, CI 0.02-0.014, P=0.000).

‘Recruit first’ trials and ‘register first’ trials change over time in Fig 3. There were 23 ‘recruit first’ trials in the past 5 years, of which 73.9% (n=17) were located in China.

**Fig 3.**
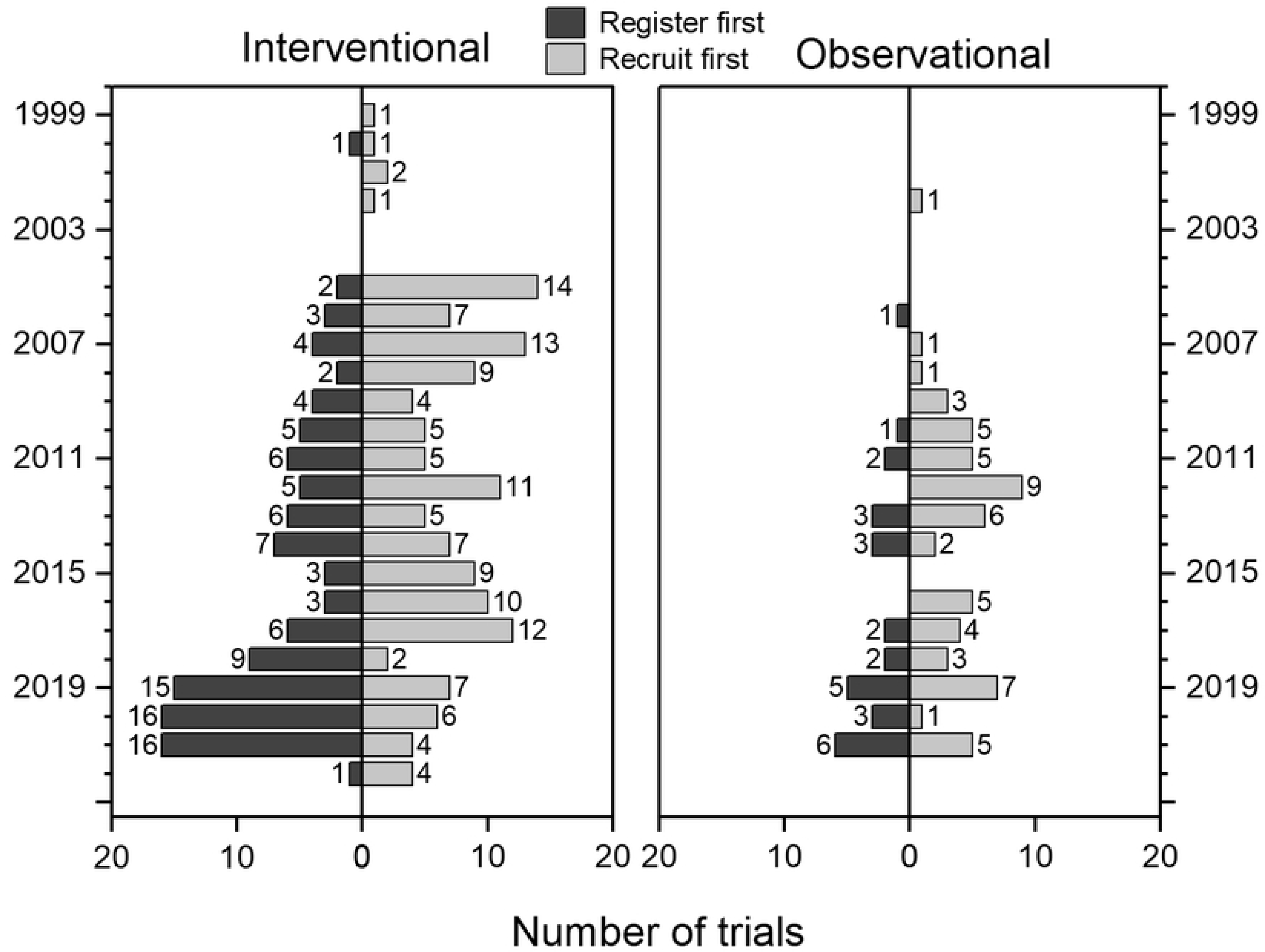
Changes in ‘register first’ trials and ‘recruit first’ trials.

The relationship between phase and study design in interventional trials involving drugs is shown in Fig 4. There were 25 Phase 1 trials (10.0%), 80 Phase 2 trials (32.1%), and 84 Phase 3 trials (33.7%). In Phase 3, 35.7% of single-arm trials (n=10) were related to immunoglobulins. There were 22 trials on immunoglobulins, so 45.5% of the immunoglobulin trials were Phase 3 single-arm trials.

**Fig 4.**
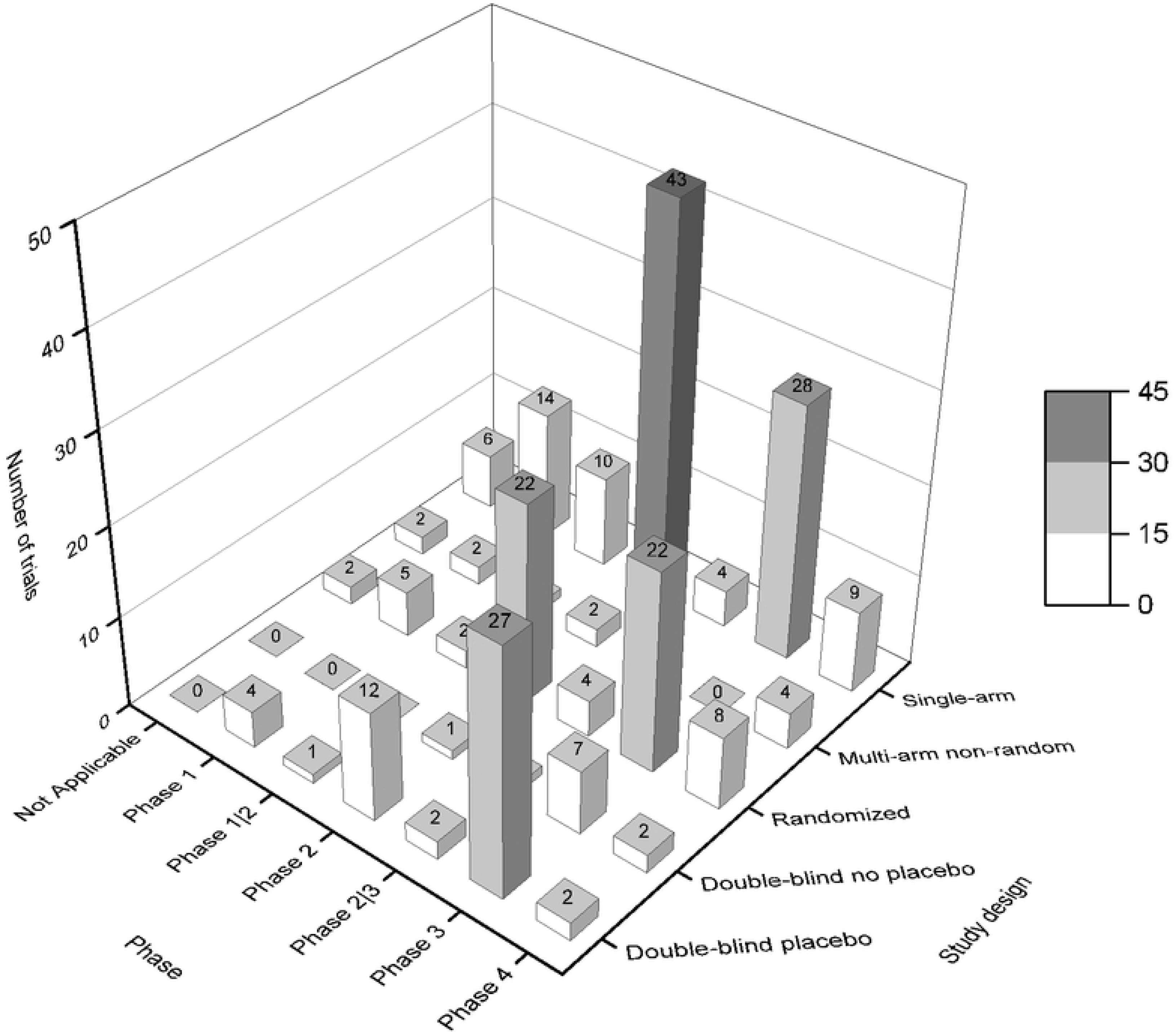
The relationship between phase and study design. The parts of the study design do not overlap. Multi-arm non-random means multi-arm non-random non-double-blind; randomized means randomized non-double-blind.

## Discussion

In the ClinicalTrials.gov database, we performed a comprehensive analysis of ITP clinical trials, half of which were in ‘Completed’ status. 55.7% of completed ITP trials published articles. Macular degeneration had 54% of the trials published but limited to interventional trials completed at the latest 2 years ago.[7] 64.4% of rheumatoid arthritis trials were published but limited to randomized controlled trials of phases 2-4 completed at the latest 2 years ago.[8] Trials with type 2 diabetes mellitus showed a publication rate of less than 40% of completed trials.[9] The reason is that the current pathogenesis of ITP has not been fully elucidated, and some patients with ITP have poor therapeutic effects, which further promotes more positive results in clinical trials.

Completed trials are required to provide their ‘basic results’ within 1 year, including participant demographics and outcome data, adverse events. Currently 28.7% of completed trials have submitted results. The upload rate of sickle cell disease and thalassemia trials in 2013 was 13%, and the upload rate of otology trials in 2019 was 21.5%.[10, 11] As time progressed, more trials began to submit results. A small number of trials with ‘Unknown status’, ‘Withdrawn’, and ‘Terminated’ status have been published, indicating that some data are inaccurate and the status of clinical trials has not been updated. Some errors in the original data were found in the process of data statistics, which have been corrected by the researchers.

In the other half, ‘Recruiting’ and ‘Unknown status’ account for the majority, indicating that some trials are in progress or the status has not been updated in time. There may be various reasons for not updating the trial status, such as changes in the actual work of the principal investigator, completed trials wanting to delay providing basic results, delaying news of trial failure. The principal investigator needs to be alerted when the status has not been updated for a certain period of time. The ‘Terminated’ and ‘Withdrawn’ trials also recommend that investigators assess whether there are enough patients enrolled and whether the sponsor agrees to sponsor the full trial before the trial begins.

During the statistical process, the researchers found some similar trials. On one hand, collaborative research can be suggested to these principal investigators who have the same idea. On the other hand, when investigators register for a clinical trial, relevant trial information will be published on ClinicalTrials.gov. Whether there are other investigators who search for relevant information and conduct similar trials, resulting in vicious competition, which is equivalent to increasing the number of patients recruited in one trial. If the treatment in the trial is effective, it may be beneficial to the patients recruited; if the treatment is suboptimal and the efficacy is poor, it may be detrimental to the inclusion of more patients. Nevertheless, it is possible to allow principal investigators to choose not to display the key content of clinical trials within a certain period of time.

The total number of trials involving China ranks first, but there are few trials conducted by pharmaceutical companies, and most of the trials focus on the use of drugs for other diseases in the treatment of ITP, combination drugs, and dose adjustment of previous drugs. Newly developed drugs that have a greater impact on the efficacy of the disease are generally discovered by companies in Switzerland, the United Kingdom, and the United States. Research on chimeric antigen receptor T cell clinical trials shows that there is a correlation between national GDP and the number of registered trials (Pearson’s coefficient of 0.92), considering that ITP clinical trials have a certain relationship with the national economy.[12] In addition to ClinicalTrials.gov, the clinical trial registries currently accredited by ICMJE include 17 centers around the world, such as EU Clinical Trials Register, Chinese Clinical Trial Registry, Clinical Trials Registry – India, etc. Therefore, some clinical trials may be registered in the regional center, resulting in the results showing that there are fewer trials in the corresponding country.

‘FDAAA 801’, the subsequent addition of ‘the Final Rule for Clinical Trials Registration and Results Information Submission’, and ‘the Checklist for Evaluating Whether a Clinical Trial or Study is an Applicable Clinical Trial’ published in the ClinicalTrials.gov database in June 2018, these legal provisions have strict registration requirements for interventional trial. For observational trial, especially retrospective study, no registration is required, so the number of observational trials is small. Fig 3 shows the sharp decrease in ‘recruit first’ trials in 2018, which is considered to be affected by ‘the Final Rule for Clinical Trials Registration and Results Information Submission’.

There are 12,000 phase 1 and 2 trials in cancer trials, but only about 1,000 phase 3 trials.[13] Statistics from October 2007 to September 2012 showed that the proportion of non-infectious disease phase 1, 2, and 3 trials accounted for 16.0%, 25.0%, and 16.6%, respectively, while all infectious diseases accounted for 17.0%, 23.1%, and 22.7%.[14] The proportion of each stage of ITP clinical trials is quite different from those of these diseases. The principal investigator of the ITP trial will treat other diseases or in combination with some drugs to treat ITP. These drugs have passed the phase 1 stage, so the number of phase 2 and 3 is significantly larger than that of phase 1. Intravenous immune globulin has a good safety profile and is expensive, so there are many single-arm intervention models.[15]

The American Society of Hematology (ASH) first published guidelines for ITP in 1996, recommending glucocorticoids, IVIg, and splenectomy as initial treatment, and accessory splenectomy, danazol, and azathioprine as complementary treatment.[16] In 2019, ASH updated the ITP guidelines, recommending glucocorticoids, IVIg as initial treatment and rituximab, eltrombopag, romiplostim, and splenectomy as complementary treatment.[17] It is the large number of clinical trials on rituximab, eltrombopag, romiplostim that have been conducted over the past 20 years or so that have led to the recommendation of these drugs and further refinement of the treatment modalities for ITP. Rituximab gave the patient remission, while eltrombopag, romiplostim gave the patient stable platelet levels.

There are 12 clinical trials focusing on glucocorticoids. The nine clinical trials focused on the difference in efficacy between prednisone and dexamethasone among glucocorticoids, while the clinical trials differed in the dose and duration of dexamethasone. 1 trial investigating the efficacy of high-dose methylprednisolone in combination with IVIG versus placebo in combination with IVIG. The results show that the combination of high-dose methylprednisolone and IVIG leads to a faster rise in platelets than IVIG alone, so the use of combination therapy is recommended in cases of life-threatening bleeding.[18]1 trial investigating the efficacy and adverse events associated with the use of 40 mg/d dexamethasone for 4 days in the treatment of patients with ITP during pregnancy. Results are not yet available. A recent trial examined whether different vitamin D levels affect the efficacy of dexamethasone in the treatment of ITP. More than thirty other trials have investigated the efficacy of other drugs in combination with glucocorticoids.

There are 12 rhTPO (Recombinan human thrombopoietin, rhTPO) related tests, and the principal investigatoris located in China. Six clinical trials with rhTPO alone, three of which studied different doses of the drug and three of which studied the efficacy and safety of the drug in pregnancy. The results of the pregnancy study showed that 23 patients (74.2%) responded, and no congenital disease or growth retardation was observed during the median follow-up of 53 weeks.[19]6 clinical trials of rhTPO in combination with other drugs, 2 of which focused on the efficacy of rhTPO in combination with rituximab for the treatment of recurrent ITP in adults. The results showed that the drug combination significantly improved CR rates, but there was no difference in long-term remission.[20]2 studies of rhTPO in combination with dexamethasone for newly diagnosed adult ITP patients. Results showed higher rates of initial response and complete complete response and longer overall duration of response in the combination group.[21]1 study combined with cyclosporine has been withdrawn.1 trial was conducted to study the efficacy of rhTPO and eltrombopta. The results showed that platelets rose more rapidly in the rhTPO group and that the most common adverse effect was fever. However, after drug withdrawal, platelet count in rhTPO group decreased to baseline within 1 week, and that in eltrombopta group decreased at the fourth week.[22]

There are 31 trials of single agent studies of eltrombopta. The clinical trials initiated in 2005 have focused on the efficacy of different doses of eltrombopta, the effects on patients with hepatic insufficiency, renal insufficiency, pregnancy and further studies on the adverse effects of eltrombopta such as eye disease. Studies in recent years have focused on the efficacy of eltrombopta in children with ITP, as well as the effect of the FcγRIIIA polymorphism on eltrombopta and the effect of eltrombopta on platelet collagen receptor glycoprotein VI. In clinical trials of eltrombopta in combination with other drugs, the effects of eltrombopta and romiplostim on platelet apoptosis have been studied in the past,[23]as well as the efficacy of the two drugs when interchanged.[24] In recent years research has focused on the use of eltrombopta in combination with other first and second line therapeutic agents for the treatment of patients with ITP, such as rituximab, rhTPO and dexamethasone.

Twenty-five trials of single-agent studies of romiplostim have focused on efficacy in patients with ITP, and on specific outcomes affecting bone marrow morphology. As romiplostim requires subcutaneous injection, clinical trials exist to study the proportion of correct use of the drug. In addition to the comparative or joint studies related to eltrombopta, there are also ITP patients who are resistant to eltrombopta treated with combination of romiplostim and danazol. In one clinical trial, romiplostim combined with low-dose rituximab and high-dose dexamethasone were used as the first-line treatment of ITP. The main regimen is Romiplostim 2mcg/Kg subcutaneously on days 1, 7, 14 and 21; Rituximab 100mg on days 1, 7, 14 and 21; Dexamethasone 40mg IV/PO on days 1-4.

A total of 21 trials of avatrombopag, hetrombopag and lusutrombopag monotherapy studies. Phase 3 clinical trials of avatrombopag showed a platelet response rate of 65.63% at day 8, with the most common adverse events being headache and contusion.[25] The results of the Phase 3 trial of hetrombopag showed a 58.9% response rate in the hetrombopag 2.5mg group and 64.3% in the hetrombopag 5mg group, with the most common adverse events being upper respiratory tract infections and urinary tract infections.[26] Relapse occurred in 88.1% of patients after stopping the drug and 74.9% achieved a platelet count ≥ 30 × 109/L at least once after reintroduction of the drug.[27]The clinical trial of lusutrombopag has been withdrawn.

Seventeen clinical trials of immunosuppressive drugs exist. A retrospective study analyzed hydroxychloroquine in combination with or without other ITP drugs for the treatment of adult patients with positive antinuclear antibodies. The results showed an overall response rate of 60%, with 75% of patients receiving hydroxychloroquine only.[28]In contrast, another trial using decitabine in refractory patients showed sustained response rates of 44.44% (20/45), 31.11% (14/45) and 20.0% (9/45) at 6 months, 12 months and 18 months respectively.[29] Common adverse events are nausea and mild fever. Decitabine is specifically administered as 3.5mg/m2 intravenously over three consecutive days, once every 4 weeks for 1 cycle, and will be used for 3 cycles. The study of dexamethasone in combination with rituximab, cyclosporine and immunoglobulin for the treatment of newly diagnosed patients started in 2016 and the results are currently unknown. The study on the treatment of chronic patients with hydroxychloroquine, vincristine and azathioprine respectively was completed in 2018, and the results are unknown.

Previous trials have shown that Alemtuzumab combined with rituximab and veltuzumab alone has a certain effect.[30, 31]

There are seven ongoing FcRn antagonist trials, three for efgartigimod, three for rozanolixizumab, and one for HBM9161. Efgartigimod can reduce IgG in the body, and one trial showed that 46% of patients treated with the drug and 25% of patients in the placebo group achieved ≥ 50×109/L at least twice. [32, 33] Trials of rozanolixizumab showed clinically relevant platelet responses (≥ 50×109/L) in more than half of patients.[34] 2 trials evaluating rozanolizumab for the treatment of persistent or chronic ITP have been terminated in September 2022 due to strategic business decisions. The trial was a multicenter, double-blind, placebo-controlled study. The primary outcome measure is the proportion of clinically significant platelet responses obtained for a sustained period of 8 weeks, with a platelet response of ≥50 × 10^9/L. An open label extension study on rozanolixizumab treatment of persistent or chronic ITP is ongoing. The primary outcome measure of the study was the occurrence of emergency adverse events. Phase 1 trials of HBM9161 showed that the drug can safely and effectively reduce IgG in Chinese subjects. [35] The dose control trial of HBM9161 double blind placebo is ongoing, and the primary outcome measure of the study is the proportion of patients with early remission at 7 weeks.

SYK inhibitors can inhibit the phagocytosis of platelets by macrophages by inhibiting SYK signalling.[36] A phase 3 randomized placebo-controlled trial showed an overall response rate of 43% for fostamatinib compared with 14% for placebo.[37] There are currently five SYK inhibitor trials underway, involving fostamatinib and other SYK inhibitors, such as: SKI-O-703, HMPL-523.

The NCT04132050 trial is a double blind placebo trial. Subjects are patients with chronic ITP and will receive 24 weeks of fostamatinib or placebo followed by a 28 week open-label period of fostamatinib treatment. The primary outcome measure for the study is the percentage of patients with a stable platelet response at week 24. A stable platelet response was defined as a platelet count ≥ 50,000/μL at least 4 out of 6 visits between week 14 and week 24.

NCT04904276 is an observational study of Fostamatinib. Subjects are ITP patients with inadequate glucocorticoid or immunoglobulin response. The primary outcome measures are change in drug dose, change in platelet count, and quality of life measures.

While a double-blind placebo trial of SKI-O-703 is underway. The study features the inclusion of a dose control group at the same time, with subjects divided into 3 treatment groups that will receive 12 weeks of treatment. In the first group, the drug dose was four 100mg SKI-O-703 capsules, twice a day; In the second group, the drug dose was two 100mg SKI-O-703 capsules combined with two placebo capsules, twice a day; The drug dose of the third group is 4 placebo, twice a day. The primary outcome measure was the percentage platelet response; and platelet response was defined as a platelet count ≥30 × 10^9/L and twice the baseline platelet count.

HMPL-523 was conducted in 2019 in a dose-escalating double-blind placebo trial. This was followed by a double-blind placebo trial in 2021 with a drug dose of 300 mg of HMPL-523 treated once daily for 24 weeks. The primary outcome measure is the durable response rate, defined as a platelet count ≥50 × 10^9 /L at least 4 out of 6 visits from week 14 to week 24.

BTK inhibitors such as rilzabrutinib inhibit B cell activation by inhibiting BCR and inhibit IgG-mediated FcγR activation.[38] Results from NCT03395210 showed that 40% of patients achieved the primary endpoint of platelet response at a median treatment day of 167.5 days.[39]

According to the previous research results of rilzabrutinib, the optimal drug dose was determined to be 400mg twice a day. At present, the double-blind placebo trial involving 224 subjects has begun. And the test subjects include some teenagers. The primary outcome measures vary according to the region. In the EU and UK, the primary outcome measure was the proportion of adult participants with a platelet count ≥50 × 10^9/L for at least 8 of the last 12 weeks of the 24 week treatment period. In other regions, the primary outcome measure was a durable platelet response during the last 6 weeks of the 24 week treatment period. Durable platelet reaction is defined as: in the last 12 weeks of the 24 week blind treatment period without rescue treatment, at least two thirds of the 8 platelet counts are ≥50 × 10^9/L. There may therefore be a partial conflict in terms of the time limit.

And 2 clinical trials of zanubrutinib are being conducted in China. The NCT05279872 clinical trial drug dose is 80mg once daily for 6 weeks of treatment. The trial is a single arm study enrolling 10 subjects. The primary outcome measure was the proportion that the platelet count remained ≥ 30 × 10 ^ 9/L at 6 weeks, and the baseline count increased at least twice. The initial dose in the NCT05214391 clinical trial was 80 mg per day. If treatment is ineffective after 4 weeks, the investigator may increase the dose to 80 mg twice daily, or a higher dose for oral maintenance. The maximum dose is 160mg twice daily. The duration of treatment is 24 weeks. The trial is a single arm study enrolling 30 subjects. The primary outcome measure is the proportion of subjects with platelet counts ≥30 × 10^9/L and 50 × 10^9/L at week 12.

In the relevant clinical trials of orelabrutinib, NCT05020288 was a single arm study involving 40 subjects. The drug dose was 50mg per day for 12 weeks. The primary outcome measure was the overall response at day 14. NCT05124028 was a single arm study that included 10 subjects. The drug dose was 50 mg per day for 6 weeks of treatment. The primary outcome measure was the proportion that the platelet count remained ≥ 30 × 10 ^ 9/L at 6 weeks, and the baseline count increased at least twice. NCT05232149 was a dose-comparison study that included 30 subjects. The study was divided into low and high dose groups, specific drug doses were not specified. The primary outcome measure was the proportion of subjects with a platelet count ≥ 50 × 10^9/L at 12 weeks.

Some single drug therapy trials are ongoing, such as CD38 antibody daratumumab, complement C1s inhibitor BIVV009/sutimlimab, oral complement bypass serine protease factor B selective inhibitor Iptacopan[40], CXCR5 antibody PF-06835375, oseltamivir, chidamide, all-trans retinoic acid, human umbilical cord mesenchymal stromal cells, fecal microbiota transplantation, several novel immunoglobulins.

There are also trials of the above mentioned drugs in combination with new drugs, such as glycyrrhetinic combined with high-dose dexamethasone, vitamin D calcitriol combined with high-dose dexamethasone, tacrolimus combined with high-dose dexamethasone, acetylcysteine combined with high-dose dexamethasone, all-trans retinoic acid in combination with low-dose rituximab, belimumab in combination with rituximab, bortezomib in combination with rituximab, diacerein with eltrombopag, terbutaline with danazol, berberine with danazol, iguratimod with danazol, the combination of atorvastatin, acetylcysteine and danazol. As a result of these clinical trials, higher treatment outcomes and a better prognosis for ITP patients will be achieved.

This study also has limitations. Although the ClinicalTrials.gov database is the largest database in the world, there are still 17 databases recognized by the ICMJE, which are not counted. Secondly, the trial information in the database is uploaded by the investigators themselves, and there are certain errors. The errors we found were also changed in the study. Third, some of the results of this article do not exclude ‘Terminated’ and ‘Withdrawn’ state trials. These trials could not be conducted due to low patient numbers or sponsorship issues. Therefore, the researchers believe that this part of the trial is also desirable.

According to the ‘World Population Prospects 2019’ released by the United Nations, the world population is predicted to reach 7,953,952,577 in 2022. Due to the novel coronavirus pneumonia epidemic, as of early July 2022, WHO data showed that there were 547,901,157 confirmed cases of coronavirus disease 2019(COVID-19), which combined with the United Nations population projections estimated that 6.8% of humans were sick. A total of 12,037,259,035 doses of the vaccine were administered, but there are many types of vaccines, and the number of vaccinated cannot be counted. Whether infection of ITP patients with severe acute respiratory disease coronavirus 2 or its variant strains aggravates the disease is unclear. However, there have been cases of ITP secondary to COVID-19. There were more men (54.8%) than women in these patients, with a median age of 63 years. [41] The mRNA COVID-19 vaccine is not the cause of ITP, and patients with ITP after vaccination may be due to the onset of their own ITP. [42] But after COVID-19 vaccination in 52 chronic ITP patients, 6 (12%) patients experienced severe platelet decline. [43] Therefore, we can consider the existence of ITP secondary to COVID-19 infection, and vaccination with COVID-19 may lead to exacerbation of ITP patients. Currently ongoing clinical trials involve types of medication efficacy, pathogenesis, diagnosis, and epidemiology. Both COVID-19 and the impact of COVID-19 vaccines on patients and clinical trials may need to be considered. In China, the above effects may be reduced due to strict management of COVID-19, but follow-up of patients may be affected.

## Conclusion

In conclusion, this study comprehensively analyzed ITP-related clinical trials in the ClinicalTrials.gov database. The results show that the difference between interventional and observational trials is mainly in the main content. The number of people included, the main trial content, the completion time, and whether the results were submitted or not will affect the publication of the trial. The publication rate of ITP clinical trials is high, but the submission rate of database results is low. Therefore, more attention should be paid to the submission of clinical trial results in the later stage. There are various drug clinical trials currently underway, and attention should be paid to the impact of COVID-19 on trials.

## Data Availability

The data underlying the results presented in the study are available from clinicaltrials.gov.

